# Anti-SARS-CoV-2 IgG antibodies are associated with reduced viral load

**DOI:** 10.1101/2020.05.22.20110551

**Authors:** Andrew Bryan, Susan L. Fink, Meghan A. Gattuso, Gregory Pepper, Anu Chaudhary, Mark H. Wener, Chihiro Morishima, Keith R. Jerome, Patrick C. Mathias, Alexander L. Greninger

## Abstract

Anti-SARS-CoV-2 antibodies have been described, but correlation with virologic outcomes is limited. Here, we find anti-SARS-CoV-2 IgG to be associated with reduced viral load. High viral loads were rare in individuals who had seroconverted. Higher viral load on admission was associated with increased 30-day mortality (OR 4.20 [95% CI: 1.62-10.86]).

## Introduction

SARS-CoV-2 is a novel coronavirus associated with high morbidity and mortality that has rapidly spread across the world. After initial concerns surrounding test characteristics, accurate serological testing for SARS-CoV-2 is increasingly becoming available in the United States. For instance, we have previously shown the Abbott Architect anti-SARS-CoV-2 nucleocapsid IgG assay to be both highly sensitive and specific for detecting prior SARS-CoV-2 infection [1]. However, data correlating these antibody results to meaningful virologic and clinical outcomes is currently lacking.

Here, we examined clinical and virologic features associated with seropositivity and seroconversion to SARS-CoV-2 in a cohort of hospitalized patients in Seattle, Washington. We specifically sought to assess whether detection of anti-SARS-CoV-2 antibodies was associated with a better prognosis, including lower viral load and reduced 30-day all-cause mortality.

## Methods

### Study population and clinical laboratory testing

Patients with positive SARS-CoV-2 RT-PCR results from nasopharyngeal swabs were identified at UW Medicine hospitals and excess serum and plasma samples were retrieved for SARS-CoV-2 antibody testing. Samples were enriched for patients that had RT-PCR results available on the same calendar date as a remnant serum or plasma sample. A total of 245 patients were identified with at least one residual serum/plasma sample and at least one clinical note available for chart review to determine days from symptom onset. The population was 40% female; age by decade was: 10-20: 0.4%, 20-29 4.9%, 30-39 6.9%, 40-49 11.0%, 50-59 17.1%, 60-69 18.8%, 70-79 21.2%, 80-89 12.2%, 90-99 7.3%. The primary clinical encounter was inpatient for 194 patients, emergency department for 39, and outpatient for 12. Eight patients were asymptomatic at the time of initial PCR result; for these patients, day of symptom onset was therefore set at the date of first positive PCR. 30-day mortality was determined by manual chart review for all patients that had either a RT-PCR result or an IgG result from the day of admission and calculated from the day of first positive PCR. The number of subjects included in the different analyses is described in the Results, as data were not available for all subjects at all time points. The study was approved under a consent waiver by the University of Washington IRB.

Anti-SARS-CoV-2 nucleocapsid IgG was determined by the Abbott Architect as previously described [1]. The manufacturer’s suggested cut-off of 1.40 was used for seropositivity. SARS-CoV-2 qRT-PCR was performed using Hologic Panther Fusion, DiaSorin Simplexa, Roche Cobas 6800 platforms, or a CDC-based laboratory developed test (LDT) [2]. Cycle threshold (Ct) values were available from the Hologic Panther Fusion and LDT assays and were treated interchangeably given their close correlation [2]. A Ct of 22 is equivalent to approximately 2,500,000 copies/mL viral transport media in these assays [3].

### Data analysis and visualization

The association between SARS-CoV-2 IgG index value and Ct was assessed using a linear mixed effects model with significance determined by restricted maximum likelihood ratio using R packages Ime4 and ImerTest [4,5]. To account for singularity, the model incorporated scaling and a weak Bayesian prior via the R package blme [6]. Multivariate logistic regression to determine association of Ct value and mortality was performed using the base R function. Visualization was performed using ggplot2 [7].

## Results

A total of 181 patients had both an Abbott Architect anti-SARS-CoV-2 nucleocapsid IgG index value and a SARS-CoV-2 PCR Ct value available from the same calendar day. Several patients had quantitative PCR and serology data available from multiple days, resulting in a total of 224 total unique patient-days. Comparison of qRT-PCR and serology data revealed only one SARS-CoV-2 seropositive individual with a simultaneous SARS-CoV-2 Ct < 22 (Figures 1A-B). While both Ct and IgG signal/index ratio increased with increasing days since symptom onset, IgG was found to be inversely correlated with SARS-CoV-2 viral load (p < 0.001). To substantiate this association at the individual patient level, we identified patients with more than three measures of SARS-CoV-2 IgG and viral load. Although the kinetics of SARS-CoV-2 IgG and viral load varied between individual patients, these parameters consistently trended together in individual patients (Figures 1C-H). Lymphocyte counts increased and inflammatory markers decreased over time in these patients (Figure S1), concomitant with a decreasing viral load (Figure 1C-H).

**Figure 1.**
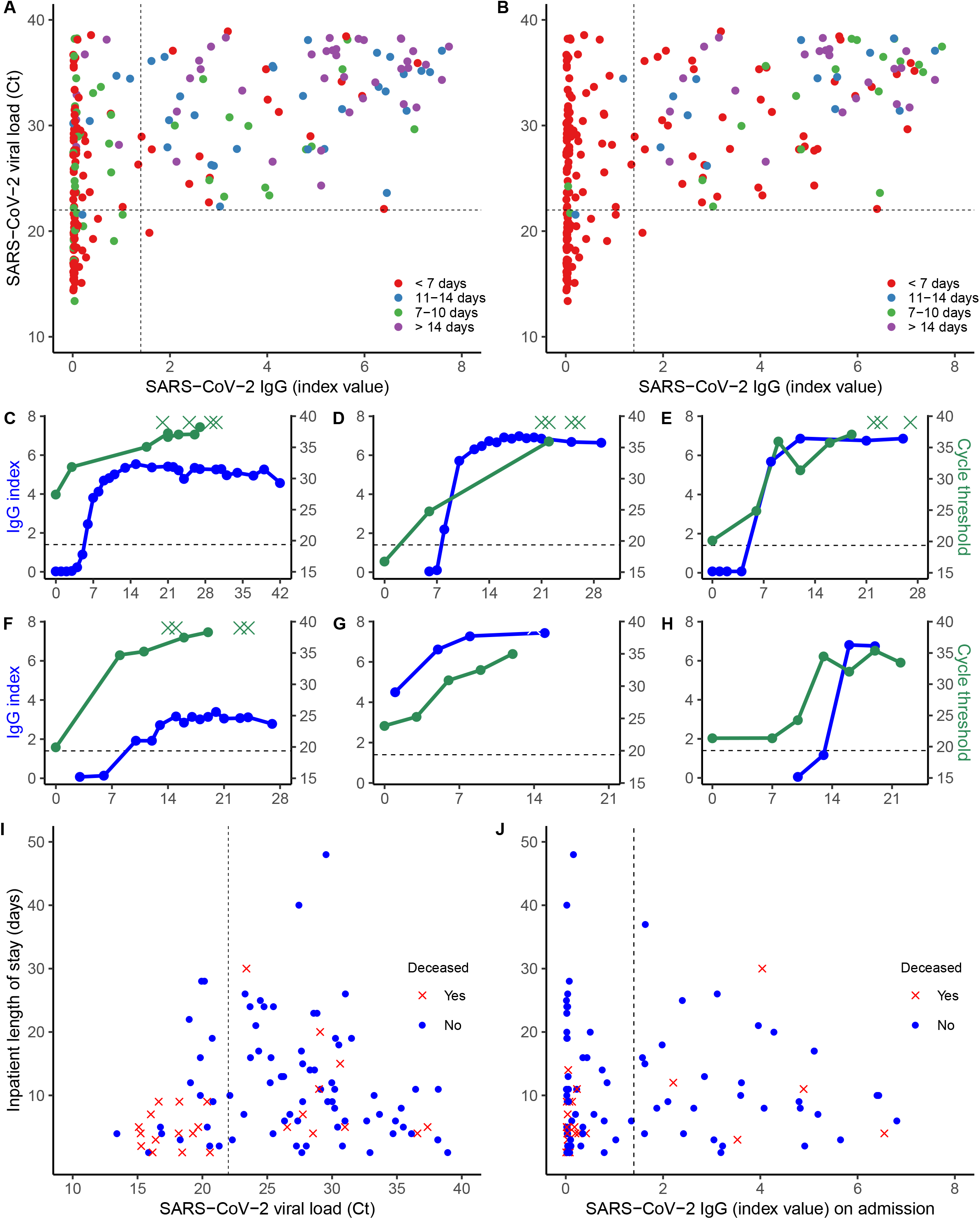
A-B) Unique patient-days with an anti-nucleocapsid IgG index value and viral load (qRT-PCR Ct value) on the same calendar day. The dashed vertical line indicates the manufacturer’s seropositivity index value cutoff of 1.40. The dashed horizontal line indicates a Ct value of 22. Colors indicate days from symptom onset (A) or days since first positive PCR (B). C-H) Six representative patients with > 3 IgG (blue) and Ct (green) results available over the course of their hospital stay. X-axis indicates days from first positive PCR. Dashed horizontal line indicates the IgG cutoff of 1.40. Green Xs indicate RT-PCR results with no nucleic acid detected. I) Patients with a Ct value available on the day of admission. Red X data points indicate patients who expired within 30 days of their first positive PCR result. The dashed vertical line indicates the manufacturer’s seropositivity threshold of 1.40. J) Patients with an IgG result available on the day of admission. Red X data points indicate patients who expired within 30 days of first positive PCR result. The dashed vertical line indicates a Ct value of 22.

To test whether seroconversion or high viral loads were associated with mortality, we examined SARS-CoV-2 viral load (n=109) and anti-SARS-CoV-2 IgG index values (n=114) on admission and 30-day all-cause mortality from day of PCR positivity (Figures 1I-J). The viral load on admission was found to be independently associated with mortality, after adjusting for SARS-CoV-2 serostatus, age, and sex (p = 0.01). Patients with a high viral load on admission (Ct < 22) had a significantly greater odds of mortality (OR 4.20 [95% CI: 1.62-10.86] compared to patients with lower viral loads (Ct > 22). Thirty-three percent of patients (38/114) seroconverted prior to admission. Seroconversion on admission trended towards lower mortality, although this relationship was not statistically significant (OR 0.43 [95% CI: 0.15-1.26]) (Figure 1J).

## Discussion

Here we demonstrate that detection of anti-SARS-CoV-2 IgG antibodies is associated with lower viral loads in COVID-19 patients. This antibody response also tracked closely with the amount of viral nucleic acid in individual patients over time. Due to the close relationships of both IgG and viral load with days since symptom onset, we could not conclude that the viral load dependence on IgG response was independent from the passage of time, but days since symptom onset was accounted for in our mixed effects model. Our data further indicate that viral load at admission is a significant independent predictor of 30-day mortality. Individuals who were SARS-CoV-2 antibody positive on admission were less than half as likely to die within 30 days, though this relationship was not statistically significant. The inability to reach statistical significance is likely due to the limited study population size (power = 0.56 for an OR of 0.5).

Our data provide support for a disease model in which the development of anti-SARS-CoV-2 immune response leads to control of the virus in humans. While our data do not directly assess the potential for ongoing immunity against future infections of SARS-CoV-2, they indicate that high viral loads almost never coexist with SARS-CoV-2 seropositivity and that antibodies are indicative of a protective anti-viral response and reduced 30-day all-cause mortality.

Our data provide further support for quantitative viral load assessment, especially on hospital admission. Our results agree with other work that has shown viral load to be associated with disease severity [8]. Currently, the FDA has only authorized reporting of qualitative results from SARS-CoV-2 qRT-PCR tests, despite nearly all tests returning some estimate of viral load. Indeed, our clinical laboratory has reported semi-quantitative results for respiratory viruses for more than a decade. Quantitative reporting of SARS-CoV-2 molecular or serologic assays would require significant modifications of existing emergency use authorizations. Our data further suggest that a cycle threshold of 22 may serve as a useful discrete cut-off for significant viral replication that is associated with mortality. We note, however, that sample and swab variability across patient populations may limit the widespread use of a discrete cutoff for quantitative RT-PCR results.

The main limitation of our study was the retrospective nature in a population enriched for hospitalized patients with acute disease. The retrospective nature precluded analyses of viral clearance and length of stay due to significant confounding factors associated with RT-PCR testing frequency during admission and patient discharge placement. Although we had insufficient sample size to perform separate analyses with patients who only presented to the emergency department or outpatient clinic, results appeared similar to the full data set (Figure S2). Our serological test detects IgG antibodies against the nucleocapsid protein of SARS-CoV-2, and the correlation between these antibodies and neutralizing antibody responses is unknown. Variability in neutralizing responses between patients not elucidated by our assay may explain some of the variation in our data set. However, neutralizing antibody assays are *in vitro* methods that may or may not be associated with clinically meaningful outcomes unless performed in concert with challenge studies. In addition, non-neutralizing antibodies may also confer protection against infection in some viral infections [9].

Our work illustrates the importance of serological testing for SARS-CoV-2 infection. The association of the presence of anti-SARS-CoV-2 nucleocapsid IgG with lower viral load indicates antibodies may serve as a biomarker for COVID-19 disease course and infectious risk of the individual to the community.

## Data Availability

Deidentified data is available from authors under reasonable request.

## Acknowledgements

The authors would like to thank Nathan Breit of the University of Washington Department of Laboratory Medicine for assistance in obtaining data and Thomas E. Grys of the Mayo Clinic in Arizona for critical review of the manuscript. We also thank the University of Washington Medical Center (UWMC) Northwest Campus clinical laboratory staff and the UWMC Clinical Immunology staff or reserving remnant serum and plasma samples from COVID-19 PCR positive patients.

## Supplemental Figures

**Figure S1.**
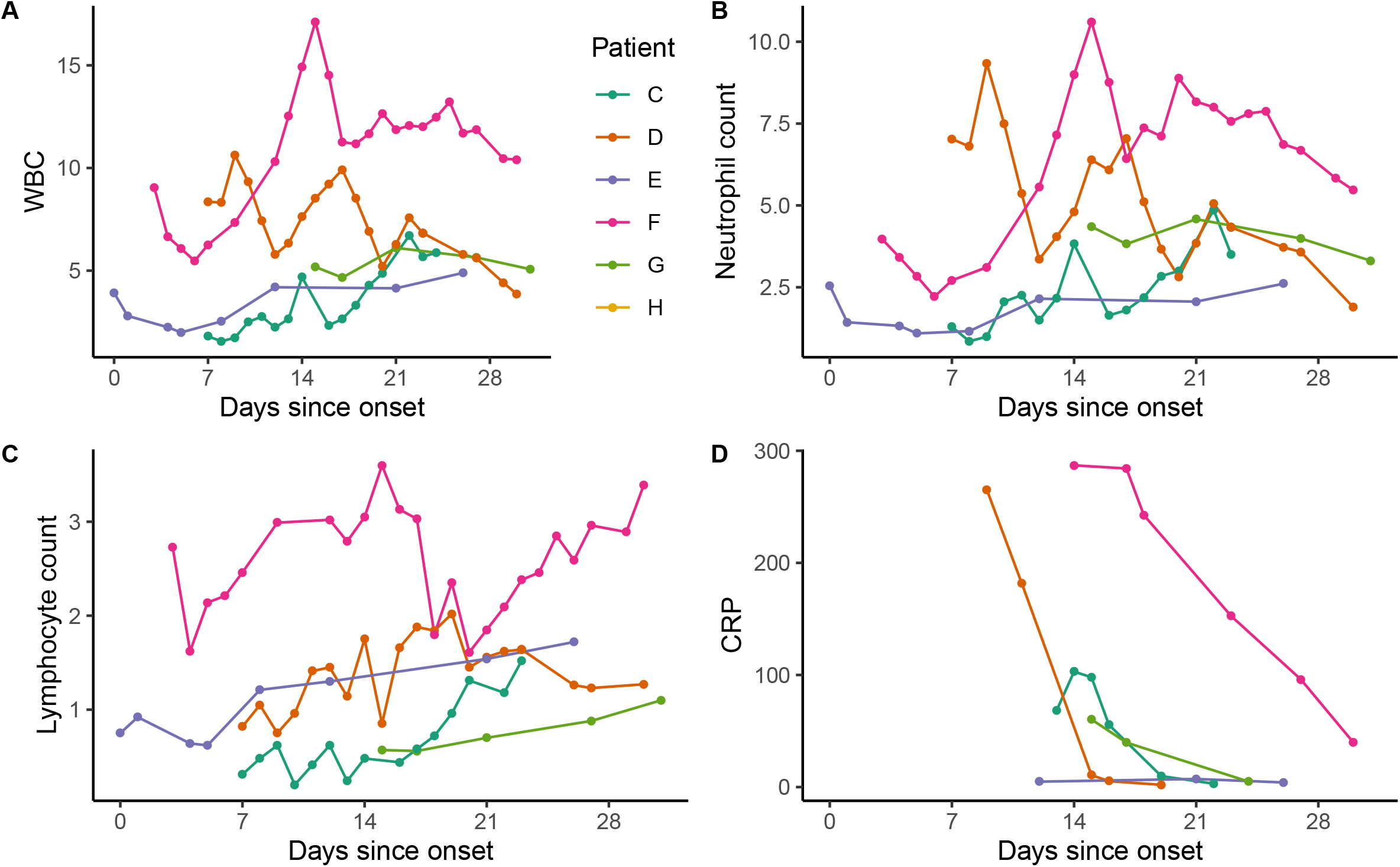
Hematology cell counts (panels A-C) or CRP by day since symptom onset for the six patients presented in Figures 1C-H. Patient H did not have cell count or CRP data available.

**Figure S2.**
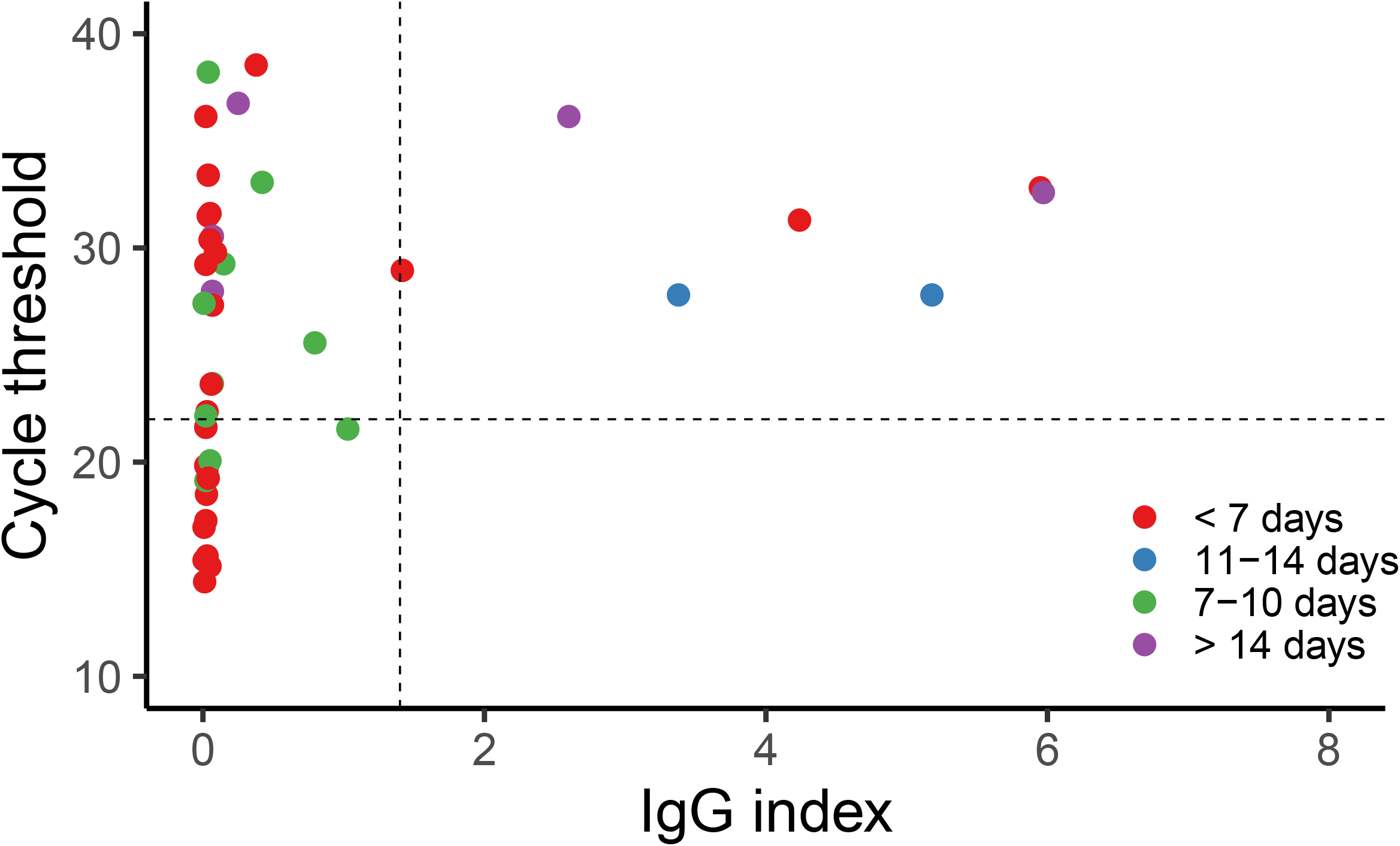
Patients who had their primary encounter as an outpatient or emergency department-only visit. Data indicate unique patient-days with an anti-nucleocapsid IgG index value and RT-PCR Ct value on the same calendar day. The dashed vertical line indicates the manufacturer’s IgG index value cutoff of 1.40. Dashed horizontal line indicates a Ct value of 22. Colors indicate days from symptom onset.

